# Capturing population differences in rates of vascular aging using a deep learning electrocardiogram algorithm: a cross-sectional study

**DOI:** 10.1101/2021.09.09.21263337

**Authors:** Ernest Diez Benavente, Francisco Lopez-Jimenez, Olena Iakunchykova, Sofia Malyutina, Alexander Kudryavtsev, Andrew Ryabikov, Paul A. Friedman, Suraj Kapa, Pablo Perel, Tom Wilsgaard, Henrik Schirmer, Taane G. Clark, Zachi I. Attia, David A. Leon

## Abstract

**Background:** Cardiovascular event rates increase with age in all populations. This is thought to be the result of multiple underlying molecular and cellular processes that lead to cumulative vascular damage. Apart from arterial stiffness based on pulse wave velocity there are few other non-invasive measures of this process of vascular aging. We have developed a potential biomarker of vascular aging using deep-learning to predict age from a standard 12-lead electrocardiogram (ECG). The difference between ECG predicted and chronological age (δ-age) can be interpreted as a measure of vascular aging.

**Methods:** We use data collected in two cross-sectional studies of adults aged 40-69 years in Norway and Russia to test the hypothesis that mean levels of δ-age, derived from a deep-learning model trained on a US population, correspond to the known large differences in cardiovascular mortality between the two countries.

**Findings:** Substantial differences were found in mean δ-age between populations: Russia-USA (+5·2 years; 0·7, 10 IQR) and Norway-USA (-2·6 years; -7, 2 IQR). These differences were only marginally explained when accounting for differences in established cardiovascular disease risk factors.

**Interpretation:** δ-age may be an important biomarker of fundamental differences in cardiovascular disease risk between populations as well as between individuals.

## Background

Fatal and non-fatal cardiovascular disease (CVD) event rates, such as stroke and myocardial infarction, increase steeply with age. This relationship is observed in all populations, independent of mean level of risk, and is the basis for regarding CVD as intimately connected with biological aging. Over the past decade much has been written about the concept of vascular aging associated with increases in factors such as arterial stiffness^1^ and the progressive build-up of atherosclerotic lesions;^2^ both of which have well-established associations with cardiovascular event rates.^3,4^ It has been postulated that there are shared molecular mechanisms of aging that promote macrovascular and microvascular pathologies^5^ including oxidative stress, mitochondrial dysfunction and chronic low-grade inflammation.^6^ However, there are few population level summary measures that attempt to quantify the cumulative overall effect of these various underlying processes on vascular aging.

We have developed a novel potential biomarker of vascular aging. This arose from work to predict age from a deep learning analysis of 12-lead ECGs.^7^ It was noted that while predicted ECG-age was highly correlated with chronological age there were deviations between the two.^7^ This led to the proposal that the direction and magnitude of the difference between ECG-age and chronological age (delta-age (δ-age)) may be a measure of relative vascular aging: if δ-age is positive, it would suggest a higher cumulative vascular damage and therefore, higher rate of vascular aging relative to the reference population from which the age algorithm was derived. This has been supported by our subsequent work that has shown δ-age to be prospectively associated with mortality,^8^ cardiovascular events^9^ and cross-sectionally with a range of established CVD risk factors including smoking and blood pressure.^10^ There is also a strong positive association between δ-age and pulse-wave velocity,^10^ an established marker of vascular stiffness, which has been regarded as one of the few direct markers of vascular aging.

In contrast to the substantial clinical and experimental work around the underlying concept of vascular aging in individuals, little attention has been given to exploring the idea that populations with different rates of CVD might be regarded as having different rates of vascular aging. It is theoretically plausible that populations with high compared to low CVD event rates may have different rates of vascular aging. To address this hypothesis we compare mean δ-age from several populations that have contrasting rates of CVD mortality and life expectancy.

## Results

We compared δ-age in two recent population-based studies of adults aged 40-69 years: the Know Your Heart (KYH; 2015-18) study in Russia and the 7 study (T7; 2015-16) in Norway. To predict ECG-age we employed a convolutional neural network (CNN) model trained on 700,000 ECGs from the Mayo Clinic (USA) (see Figure 1a). By definition the predicted age for the Russian and Norwegian populations was relative to the reference clinical population in the USA. Rates of CVD mortality in Russia are over seven times greater than in Norway in the 40-69 year age group. Given these differences in CVD mortality we hypothesised that the mean difference between ECG predicted and chronological age (δ-age) for Russia would be positive, while the equivalent mean value of δ-aged for Norway would be negative. We also investigated how far any differences in mean delta-age between the Russian and Norwegian study populations could be explained by differences in known CVD risk factors.

**Figure 1.**
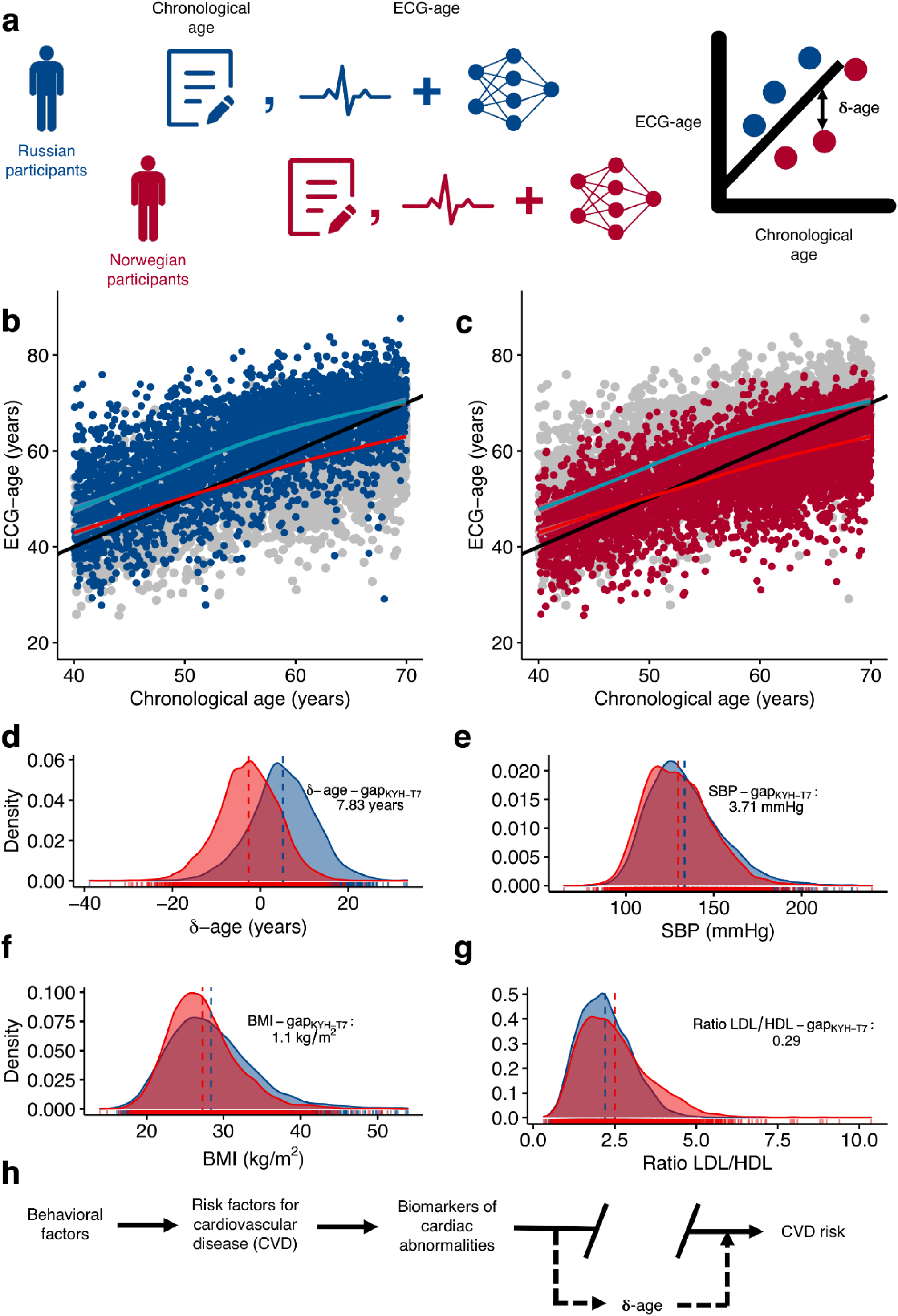
A novel digital ECG biomarker of vascular aging for population level assessment of CVD risk. **a)** Schematic followed to obtain ECG-age and form participants of both Tromsø 7 (T7) and Know Your Heart (KYH) studies, chronological age was obtained via questionnaire data and ECG-age was obtained by performing an, at least, ten second resting 12-lead ECG and processing the ECG raw data through a CNN model trained on a USA clinical population to predict age. δ-age was obtained by subtracting the chronologic age to the ECG-age. **b)** Scatterplot of ECG-age versus chronological age for 3,487 participants form KYH (Russia) in blue and 4,704 T7 participants from Norway in grey. Locally weighted smooth line was presented in light blue for KYH and light red for Norway. **c)** Scatterplot of ECG-age versus chronological age, in this panel dark red dots represent T7 participants (Norway) and grey dots represent KYH participants (Russia). This panel presents the same data as panel **b)** with different colouring for clarity. **d)** Density plot of δ-age for Russia and Norway, δ-age-gap is not adjusted. **e)** Density plot of systolic blood pressure (SBP) for Russia and Norway, SBP-gap is not adjusted. **f)** Density plot of body mass index (BMI) for Russia and Norway, BMI-gap is not adjusted. **g)** Density plot of ratio of low-density lipoprotein (LDL) cholesterol over high density lipoprotein (HDL) cholesterol for Russia and Norway, Ratio LDL/HDL-gap is not adjusted. **h)** Proposed theoretical framework depicting δ-age in the causal pathway of cardiovascular disease (CVD) as a proximal biomarker of CVD risk which stores information about both behavioural and risk factors as an accumulated exposure over time. Note: The line of identity in panels **b)** and **c)** can be seen as a proxy for the American population, representing an average δ-age of +0·44 years.^7^

Digitised ECGs were available for 5,459 T7 participants and 3,973 KYH participants aged between 40 and 69 years. After exclusion of records with missing data (N_T7_=74, N_KYH_=486) a final set of 5,385 T7 and 3,487 KYH participants were retained for analysis. The average age of the participants was 59 ± 8·3 years (54·7% female) for T7 and 56 ± 8·6 years (58·4% female) for KYH (Table S1). Mean δ-age was +5·2 years (0·7, 10 IQR) for the KYH (Russia) study population, (Figure 1b) and -2·6 (-7, 2 IQR) for T7 (Norway) study population (Figure 1c). The mean difference in δ-age between KYH and T7 studies (δ-age gap_KYH-T7_) was +7·8 years (95% CI: 7·5, 8·1) showing that the Russian KYH participants had an average ECG-age that was nearly 8 years older than Norwegian T7 participants (Table 1). Having adjusted for both chronological age and sex, the gap in δ-age between the two study populations (KYH-T7) was 7·0 years (95% CI: 6·7, 7·2; *p*<0·0001).

**Table 1.** Table of explanatory linear models for the absolute difference in δ-age between the T7 and KYH studies (δ-age-gap_KYH-T7_). The second column (δ-age-gap_KYH-T7_ explained) is the difference between the unadjusted δ-age-gap_KYH-T7_ and the δ-age-gap_KYH-T7_ of each adjusted model, reported as a percentage of the unadjusted δ-age-gap_KYH-T7_.

| Model | δ-age-gap<sub>KYH-T7</sub><br>(years) | 95% CI | p-value | δ-age-gap<sub>KYH-T7</sub><br>explained (%) |
| --- | --- | --- | --- | --- |
| Model 1: Not adjusted | 7.8 | [7.5, 8.1] | <0.0001 | - |
| Model 2: Chronological age and sex | 7 | [6.7, 7.2] | <0.0001 | 7.8 |
| Model 3: Model 2, SBP, BMI, Smoking and Ratio LDL/HDL adjusted | 6.5 | [6.2, 6.8] | <0.0001 | 16.5 |
| Model 4: Model 3, DBP, Pulse rate, HbA1c, Education | 6 | [5.7, 6.3] | <0.0001 | 23.2 |
| Model 5: Model 4, history of hypertension and history of diabetes adjusted | 5.8 | [5.5, 6.2] | <0.0001 | 25.8 |

The *loess* smoothed fitted line for KYH (Figure 1b) was above the line of identity at all ages. For T7 (Figure 1c) the fitted line was below the line of identity from age 50 years. However, the δ-age gap_KYH-T7_ was positive across the entire population, as shown by the fitted line for KYH being above that of T7 for the entire range of chronological age shown, the gap being slightly larger at older ages. There were small differences in δ-age between men and women in KYH (women were predicted 0.3 years younger on average, *p*=0·28) and T7 (0·3 years, *p*=0·15).

The distribution of δ-age in the two studies (Figure 1d) showed a marked displacement of the T7 population to the left of the KYH population. In contrast there is far less separation of the distribution of established CVD predictors in the two study populations (Figures 1e-g), although differences in the mean values are slightly higher in KYH than in T7. The degree of separation of these various distributions may be quantified in terms of a dissimilarity index that does not depend upon the shape of the distribution.^11^ This distribution has a range from zero (identical) to 1 (no overlap). The dissimilarity index for δ-age was 0·61 (95% CI: 0·59 – 0·62), compared with 0·13 (95% CI: 0·11 – 0·16) for SBP, 0·21 (95% CI: 0·19 – 0·23) for BMI, and 0·20 (95% CI: 0·18 – 0·22) for LDL/HDL ratio, reinforcing the notion of δ-age having increased power to cross-sectionally differentiate between populations presenting higher CVD risk than established CVD risk factors.

The established CVD risk factors SBP, BMI, ratio LDL/HDL and smoking were associated with δ-age in both studies (Table S2). Adjustment for these cross-sectionally measured risk factors along with sex and age reduced the gap in δ-age between studies to 6·5 years (95% CI: 6·2, 6·8; *p*<0·0001) (Table 1).

Finally, we adjusted for a larger set of established CVD-related risk factors which also showed significant differences between KYH and T7 (Table S1). These were diastolic blood pressure, pulse rate, haemoglobin A1c (HbA1c) and education level. This further adjustment reduced the gap in δ-age between studies to six years difference (95% CI: 5·7, 6·3; *p*<0·0001), with a slight reduction in the gap when further adjusting for history of hypertension and diabetes and the final model accounted for 25·8% of the gap in δ-age between studies (Table 1).

## Discussion

These results confirm our hypothesis that mean difference between predicted age and chronological age (δ-age) in the study populations in Russia was substantially different to the difference in the Norwegian study, consistent with the much higher CVD mortality rates in Russia compared to Norway. Moreover, relative to the US clinical reference population mean δ-age was higher in KYH (Russia) but lower in T7 (Norway), in line with the intermediate position of CVD mortality in the US between Russia and Norway. However, while the reference population in the US was from a clinical series, the Russian and Norwegian studies were population-based studies. As discussed elsewhere, CVD mortality rates in the Norwegian and Russian cities from which the study participants were drawn were very close to their respective national averages.^12^

Showing that mean δ-age is so much higher in the Russian compared to Norwegian study population strongly suggests that δ-age may be a robust biomarker of relative vascular aging between populations as well as being a predictor of mortality and associated with CVD risk factors within populations. While CVD mortality is not the only determinant of differences in life expectancy between countries, it is notable that the difference in δ-age between Russia and Norway (7·8 years) is similar to the difference in life expectancy at birth between the two countries (8·8 years in 2016).

Adjustment for a wide range of CVD risk factors attenuated the difference between KYH and T7 by just under 20% (from 6·95 to 5·64 years). This is consistent with recent work that suggests that a core set of established CVD risk factors were able to explain only a quarter of the observed differences in CVD mortality between Russia and Norway at ages 40-69 years.^13^ There are a number of explanations for the inability of cross-sectional risk factor profiles to explain differences in CVD mortality *per se* that may also apply to the moderate attenuation of differences in δ-age. These explanations include within person variability in the risk factor levels from single cross-sectional measures such that they provide inadequate assessments of long-term risk factor levels integrated over the life course.^13^

As we have demonstrated for a selected set of established CVD risk factors, there is far greater overlap in their distributions between the Russian and Norwegian studies compared with that for δ-age. It is therefore not that surprising that they have limited scope to explain differences in δ-age between the populations.

The appreciable difference between T7 and KYH in mean δ-age is reflected in the shift to the left of the whole T7 distribution of δ-age relative to KYH. How can we interpret this? In contrast to established predictive risk factors, the information from the ECG captured by the CNN generated algorithm for predicted age could be regarded as a direct expression of the accumulation of age-related damage or impairment to the vascular tree and its effects on cardiac structure and function. From this perspective, δ-age may reflect a net integrated effect on vascular aging that is the result of many different processes and cumulative exposures to both known and unknown risk factors. In this sense this integrated measure of age-related changes in cardiac structure and function may be far more proximal to the ultimate causes of CVD events and mortality than are single or even multiple measures of predictive risk factors.

The specific ECG features used by the δ-age algorithm remain unclear. Recent advances have been able to highlight the segments of an ECG that a CNN algorithm uses to make predictions^14^. However, the unresolved functioning of the algorithm does not prevent the objective evaluation of its potential utility, especially when studying CVD risk at a population level, where the benefit obtained from the ability to measure CVD risk trends is likely to outweigh any biases at the individual level which could have a more direct effect if this measure was used in a clinical setting. Through the use of epidemiological approaches, we have identified the ability of δ-age to replicate between population patterns of CVD risk and mortality suggesting that it captures fundamental average differences in cardiac structure/function within populations.

We believe this biomarker may enable us to close some of the knowledge gap between established risk factors and predictors and proximal measures of CVD risk at a population level (Figure 1h). Further studies would require population level data from more diverse populations, in order to ascertain the ability of δ-age to reproduce country-level CVD event trends as well as using cohorts with both raw ECG and CVD events follow-up data. Additionally, ECG signals could be used outcomes in GWAS studies to determine potential genetic contributions to δ-age and provide insights into new biological pathways relevant to vascular aging. Finally, the use of this marker if further validated may enable novel studies of CVD risk in low- and middle-income countries where resources for more complex phenotyping are scarce.

## Materials and Methods

We used data obtained in two population studies, the Tromsø 7 (T7), the 7th survey of the Tromsø Study in Tromsø, Norway^15^ and the Know Your Heart (KYH) study, a cross-sectional survey of a random sample of the population of two Russian cities: Arkhangelsk and Novosibirsk.^12^ A total of 21,083 men and women aged 40-99 years were recruited from the general population, as described in detail elsewhere.^16^ The KYH study participants were comprised of 4,542 men and women aged 35-69 years recruited from the general population between 2015 and 2018 as described in detail elsewhere.^12^

For T7 65% of those contacted and invited to take part attended the health check. A randomly selected set of 13,028 participants (9,925 randomly selected and 3,103 selected based on participation in previous rounds) were invited for a second visit where a resting 10 seconds digital 12-lead ECG was obtained using the Schiller device AT104 PC and the raw ECG signal was stored digitally. For KYH of those contacted and invited to take part 47% attended the health check. A resting 10-25 seconds digital 12-lead ECG was obtained using the Cardiax device (IMED Ltd, Hungary) and the raw ECG signal was stored digitally.

A convolutional neural network (CNN) model using Keras with a Tensorflow (Google, Mountain View, CA) backend was previously developed and validated. A total of 774,783 unique subjects with ECGs were used to develop the neural network: 399,750 in the training set, 99,977 in the internal validation set and 275,056 ECGs in the holdout testing set. The network contained stacked blocks of convolutional, max pooling, and batch normalization.^17^ A detailed description of the network is described in our previously published paper.^7^

We used linear regression models to assess association of the risk factors as exposures using δage as the outcome. All models were adjusted for a priori confounders including chronologic age and sex. True chronologic age was included in every model to remove the effects of any potential correlation between δ-age and chronologic age. A test for trend for smoking was performed by converting the ordered categories of number of cigarettes smoked into integer values. Basic models including one risk factor at a time were further adjusted for potential confounders.

Density plots were generated for δ-age, SBP, BMI and Ratio LDL/HDL. In order to assess the overlap of the density plots for each variable in the populations we used the R package *overlapping* with a 100 bootstraps per variable.^11,18^

For further details see Supplementary Methods.

## Data Availability

The data from the Know Your Heart (Russia) study be requested through the study metadata portal (https://metadata.knowyourheart.science/) upon registration. Registered users are able to browse the variable-level metadata for the Know Your Heart study; however, the site does not provide access to the data themselves. Bona fide researchers may apply to the study steering group for an anonymised subset of the data. The primary criteria used by the steering group are scientific coherence of the proposed use of the data and the match between the stated aims of the proposed research and the variables requested. This means that applicants need to justify the sets of variables they wish to have. In some instances, the research topic of the application may be close to or replicates ongoing research either within the core team or by previous applicants. In these cases, this overlap would be communicated to the applicants with a suggestion for the way forward. More information can be found in the ‘User Guide’ document on the website. Guidance on requesting data from the Tromsø study is given at https://uit.no/research/tromsostudy/project?pid=709148

https://metadata.knowyourheart.science/

## Acknowledgments

We would like to thank all the team that undertook the field work in the Know Your Heart and Tromsø 7 studies and all the participants and George Davey Smith for helpful comments on an earlier draft. Part of DAL’s contribution was undertaken within the framework of the Basic Research Program of the National Research University Higher School of Economics.

## Author Contributions

EDB, PP, DAL and FJL conceived the study, DAL, TGC, HS and TW designed the study, EDB performed the statistical analysis under the supervision of DAL and TGC. ZIA contributed to the analysis and interpretation of the machine learning analysis under the supervision of FJL, SK and PAF. AK contributed to the acquisition, analysis and interpretation of the data from Arkhangelsk in the Know Your Heart Study, SM and AR contributed to the acquisition, analysis and interpretation of data in the Know your Heart study, HS, OY and TW contributed to the acquisition, analysis and interpretation of data in the Tromsø 7 study. EDB wrote the first draft of the manuscript. All authors commented and edited on various versions of the draft manuscripts and gave final approval to the submitted version and agree to be accountable for all aspects of work ensuring integrity and accuracy.

## References

1 Donato AJ, Machin DR, Lesniewski LA. Mechanisms of Dysfunction in the Aging Vasculature and Role in Age-Related Disease. Circ Res 2018; 123: 825–48.

2 de Groot E, Hovingh GK, Wiegman A, et al. Measurement of arterial wall thickness as a surrogate marker for atherosclerosis. Circulation 2004; 109: III33–8.

3 Zhong Q, Hu M-J, Cui Y-J, et al. Carotid-Femoral Pulse Wave Velocity in the Prediction of Cardiovascular Events and Mortality: An Updated Systematic Review and Meta-Analysis. Angiology 2018; 69: 617–29.

4 Bamberg F, Sommer WH, Hoffmann V, et al. Meta-analysis and systematic review of the long-term predictive value of assessment of coronary atherosclerosis by contrast-enhanced coronary computed tomography angiography. J Am Coll Cardiol 2011; 57: 2426–36.

5 Ungvari Z, Tarantini S, Sorond F, Merkely B, Csiszar A. Mechanisms of Vascular Aging, A Geroscience Perspective: JACC Focus Seminar. J Am Coll Cardiol 2020; 75: 931–41.

6 Zoltan U, Stefano T, J. Da, Veronica G, Anna C. Mechanisms of Vascular Aging. Circ Res 2018; 123: 849–67.

7 Attia ZI, Friedman PA, Noseworthy PA, et al. Age and Sex Estimation Using Artificial Intelligence From Standard 12-Lead ECGs. Circ Arrhythmia Electrophysiol 2019; 12: e007284.

8 Adetola L, Jose M-I, Michal S, et al. ECG-derived age and survival: validationg the concept of physiologic age detected by ECG using artificial inteligence. J Am Coll Cardiol 2020; 75: 3469.

9 Medina-Inojosa J, Ladejobi A, Attia Z, et al. The association of artificial intelligence-enabled electrocardiogram-derived age (physiologic age) with atherosclerotic cardiovascular events in the community. Eur Heart J 2020; 41. DOI:10.1093/ehjci/ehaa946.2905.

10 Diez Benavente E, Jimenez-Lopez F, Attia ZI, et al. Studying accelerated cardiovascular ageing in Russian adults through a novel deep-learning ECG biomarker [version 1; peer review: awaiting peer review]. Wellcome Open Res 2021; 6. DOI:10.12688/wellcomeopenres.16499.1.

11 Pastore M, Calcagní A. Measuring Distribution Similarities Between Samples: A Distribution-Free Overlapping Index. Front. Psychol.. 2019; 10: 1089.

12 Cook S, Malyutina S, Kudryavtsev A V, et al. Know Your Heart: Rationale, design and conduct of a cross-sectional study of cardiovascular structure, function and risk factors in 4500 men and women aged 35-69 years from two Russian cities, 2015-18 [version 3; peer review: 3 approved]. Wellcome Open Res 2018; 3. DOI:10.12688/wellcomeopenres.14619.3.

13 Trias-Llimós S, Pennells L, Tverdal A, et al. Quantifying the contribution of established risk factors to cardiovascular mortality differences between Russia and Norway. Sci Rep 2020; 10: 20796.

14 van de Leur RR, Blom LJ, Gavves E, et al. Automatic Triage of 12-Lead ECGs Using Deep Convolutional Neural Networks. J Am Heart Assoc 2020; 9: e015138.

15 Jacobsen BK, Eggen AE, Mathiesen EB, Wilsgaard T, Njølstad I. Cohort profile: the Tromso Study. Int J Epidemiol 2012; 41: 961–7.

16 Wynn R, Oyeyemi SO, Budrionis A, Marco-Ruiz L, Yigzaw KY, Bellika JG. Electronic Health Use in a Representative Sample of 18,497 Respondents in Norway (The Seventh Tromsø Study - Part 1): Population-Based Questionnaire Study. JMIR Med Inf 2020; 8: e13106.

17 Ioffe S, Szegedy C. Batch Normalization: Accelerating Deep Network Training by Reducing Internal Covariate Shift. In: Bach F, Blei D, eds. Proceedings of the 32nd International Conference on Machine Learning. Lille, France: PMLR, 2015: 448–56.

18 Inman HF, Bradley EL. The overlapping coefficient as a measure of agreement between probability distributions and point estimation of the overlap of two normal densities. Commun Stat - Theory Methods 1989; 18: 3851–74.

